# Covid19db – An online database of trials of medicinal products to prevent or treat COVID-19, with a specific focus on drug repurposing

**DOI:** 10.1101/2020.05.27.20114371

**Authors:** Pan Pantziarka, Liese Vandeborne, Lydie Meheus, Gauthier Bouche

## Abstract

**Background:** The global pandemic caused by SARS-CoV-2 virus has prompted an unprecedented international effort to seek medicines for prevention and treatment of infection. Drug repurposing has played a key part in this response. The rapid increase in trial activity has raised questions about efficiency and lack of coordination. Our objective was to develop a user-friendly, open access database to monitor and rapidly identify trials of medicinal products.

**Methods:** Using the US clinicaltrials.gov (NCT) registry, the EU Clinical Trials Register (EUCTR) and the WHO International Clinical Trials Registry Platform (WHO ICTRP), we identified all COVID-19 trials of medicinal products. Trials that were out of scope and duplicates were excluded. A manual encoding was performed to ascertain key information (e.g. trial aim, type of intervention etc). The database, Covid19db, is published online at: http://www.redo-project.org/covid19db/.

**Results:** Descriptive statistics of the database from April 4th 2020 through to August 18th show an increase from 186 to 1618 trials, or an average of 10.5 new trials registered per day. Over this period, the proportion of trials including a repurposing arm decreased slightly (from a maximum of 75% to 64% at the end of the covered period) as did the proportion of trials aiming to prevent infection (from a maximum of 16% to 13%). The most popular trial intervention is hydroxychloroquine (212 trials), followed by azithromycin (64 trials), tocilizumab, favipiravir and chloroquine (145 trials). Total planned enrolment is 1064556 participants as of 18^th^ August 2020.

**Conclusions:** we have developed an open access and regularly updated tool to monitor clinical trials of medicinal products to prevent or treat infection by SARS-CoV-2 globally. Our analysis shows a high number of ‘me-too’ trials, in particular for some repurposed drugs, such as hydroxychloroquine, azithromycin and tocilizumab, substantiating calls for better coordination and better use of trial resources.

## Background

The global pandemic caused by the SARS-CoV-2 virus has led to a rapid and sustained response in terms of clinical trials investigating strategies for prevention of infection and treatment of coronavirus disease 2019 (COVID-19) caused by the infection. The rapid proliferation of trials has been unprecedented and has, inevitably, raised concerns about efficiency, duplication of effort, questionable designs and the choices of interventional and comparator agents [1,2]. Tracking the number and range of clinical trials has also been complicated by the sheer volume of trials – many of them non-interventional - and the issue of duplicate registrations. The latter occurs when the same trial is entered in multiple registries. In such cases different identifiers and other information may be used, thereby masking the fact that the same trial is being referred to.

Many of the interventional trials have adopted a drug repurposing strategy [3] – that is rather than use novel drugs or agents developed specifically for COVID-19 they have selected drugs already licensed and approved for other medical conditions [4]. Anecdotal evidence suggested that a large proportion of COVID-19 trials were using repurposed drugs, but confirmation of the actual prevalence of repurposed drugs had not been done in a systematic way. In addition to the issue of duplicated trial registrations, a further complication arose because repurposed drugs were being used as standard of care in some trials and as investigational agents in others, as illustrated by the use of the anti-viral combination of lopinavir/ritonavir in 2 randomized trials from China [5,6].

Given our pre-existing interests in drug repurposing, and the need to clarify the landscape of investigational agents in COVID-19-related trials we have produced a curated database of interventional COVID-19 clinical trials testing medicinal products. This database, which we have called Covid19db, has been released as open access on the same domain as an existing drug repurposing database ReDO_db [7]. The URL for the database is http://www.redo-project.org/covid19db/.

The objective of this report is to present the methodology used to identify all trials of medicinal products and to build the database, and to describe the characteristics of the trials with a specific focus on repurposed drugs.

## Methods

We define medicinal products as any substance, or combination of substances, intended to treat or prevent COVID-19. In addition to drugs it includes vaccines and cell- and blood-based products such as convalescent plasma and stem cell therapies. It does not include trials of medical procedures, equipment, diagnostic tools, psychological interventions or traditional Chinese medicine (TCM).

We used the following clinical trial registries as source data: the US clinicaltrials.gov (NCT) registry, the EU Clinical Trials Register (EUCTR) and the WHO International Clinical Trials Registry Platform (WHO ICTRP). ‘covid-19’ is now a term in the database ontology of all three platforms and therefore this was used as the search term for each of these registries.

The ClinicalTrials.gov API (https://www.clinicaltrials.gov/ct2/resources/download) was used to download clinical trial records, in the XML schema defined by the API, as tab-separated values, with one clinical trial record corresponding to one row in the downloaded data. Query parameters were used to include Interventional trials only, all trial phases and all recruitment statuses. The data download and processing was performed using custom code in an Excel workbook, which acted as the master file for the database. The process was designed to support an iterative workflow so that repeated queries could be performed to incorporate both new trial registrations and amendments to existing trials.

The EU Clinical Trials Registry does not provide an API and therefore a web spider was used to download a query page containing a list of COVID-19 clinical trials (https://www.clinicaltrialsregister.eu/ctr-search/search?query=covid-19). From this page a list of URLs is constructed containing the web pages of trial protocols, which could then be spidered and relevant data fields extracted from the downloaded HTML content. Where a number of national protocols exist for a trial the UK version, for language reasons, was used as the data source if it existed, otherwise the first protocol in the list was used. A mapping to the NCT XML schema was used to construct a data record that aligned with the NCT data records – for example trial registration numbers, title, recruitment status etc. The downloaded EUCTR data is thus aligned with the NCT data in the master file. A similar iterative workflow means that updates and additions can update the master file on an ad hoc basis.

The WHO ITCRP search portal came under sustained heavy traffic during the pandemic was therefore made inaccessible for non-WHO users. However, COVID-19 trials were made available in a series of Excel workbooks which were periodically released publicly. The data fields in these files were mapped to the NCT XML schema such that it could be aligned with the NCT and EUCTR data. All types of clinical trial (i.e. including non-interventional trials) were included in these WHO data extracts and therefore a filtering process was used to remove these trials. Similarly, trials already listed in NCT or EUCTR were also excluded as it was assumed that they would be sourced directly from the appropriate registry.

Data from all three sources was therefore combined into a single data table which constituted the input data for the generation of the Covid19db. Each trial in this input data set was manually assessed by one or more of the authors to ascertain the following information:

- Main aim of the trial: One of: Prevention, Therapeutic, Diagnostics, Supportive Care or Other
- Type: The type of intervention in the trial. One or more of: Drug Repurposing, Vaccine, Shelved drug or new molecular entity (NME), Cell or blood-based product, traditional Chinese medicine (TCM), Other, N/A
- Controlled (having a control arm): Yes/No
- Multi-arm (having more than one experimental arm): Yes/No
- Country of the principal investigator (PI). Single selection from drop-down list of countries. For international trials, only the country of the primary sponsor was selected. Therefore there is likely to be an underestimate of the number of trials running in each country.
- Population: One or more of: Exposed individuals (family members, health care professionals), General Population, Mild/Moderate cases, Severe cases, Other
- Drug International Non-proprietary Name (INN): One or more drugs used in the investigational arms of the trial. Drugs used on the comparison arms of trials, even if considered investigational in other trials, were not included in the coding. For example, in some trials hydroxychloroquine was considered as the standard of care and administered to all patients, therefore it was not listed in the Drug INN field for those trials.
- Primary end-point: One of: Efficacy - WHO scale (or adaptation of it), Efficacy - Clinical improvement (except WHO scale), Efficacy – Mortality, Efficacy - Oxygen parameters, Efficacy - Viral load, Efficacy - Infection rate (prevention), Efficacy - Other efficacy, Safety, Other

Comment fields were used to record additional information. A screenshot of the dialog box used to display a clinical trial record and to enter the manually assessed data is shown in Figure 1.

**Figure 1.**
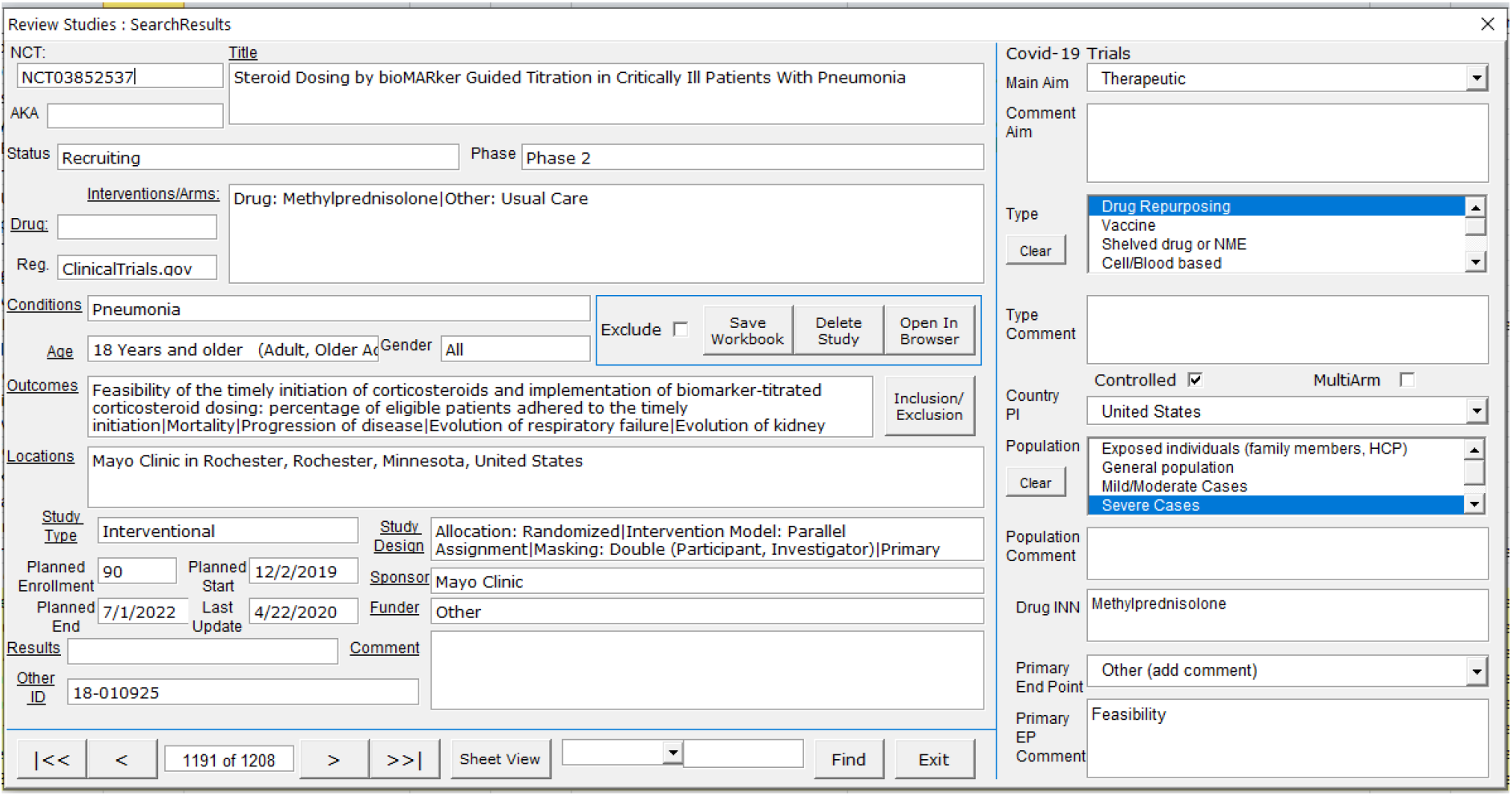
Data coding dialog screen.

In addition to entering the coding for each trial, the review process was also used to exclude trials deemed out of scope. Out of scope trials included trials incorrectly listed as interventional but actually observational only and duplicate records. Duplicate records were largely the result of multiple registrations, for example in NCT and EUCTR. In some cases identification was straightforward, for example when the registration listed other identifiers which included registration numbers for other registries. In other cases identification of duplicates was more problematic and involved comparing interventions, planned enrolment, institutions etc.

All trials which were classified as in-scope and appropriately coded were included in the analysis and output dataset. This is the set of clinical trial records which was termed Covid19db and exported for analysis and presentation online.

The online version of the database was designed to provide the most straightforward access to the data. It is presented as a simple table on a HTML page, with interactive filtering on trial ID, drug, country or type of intervention. The data is also available as a tab-separated values file so that it can be downloaded and used for additional analysis or searching. Both the online and TSV formats include hyperlinks to the original trial registration web pages.

## Results

The first public release of Covid19db was on April 7 2020. At that point the database contained 211 interventional trials, of which 186 were testing medicinal interventions and all but 12 trials were sourced from NCT or EUCTR. Analysis of the rate of increase is complicated by the fact that the data from Chinese registered trials included in the ICTRP stream was not fully integrated until April 21^st^. However, we can observe the rate of increase in registered trials by looking at the NCT and EUCTR trials in the database, as shown in Figure 2 (from April 4^th^ to August 18^th^). On average, 10.5 new trials were registered each day in that period, increasing from 186 trials on April 4th to 1618 trials on August 18th.

**Figure 2.**
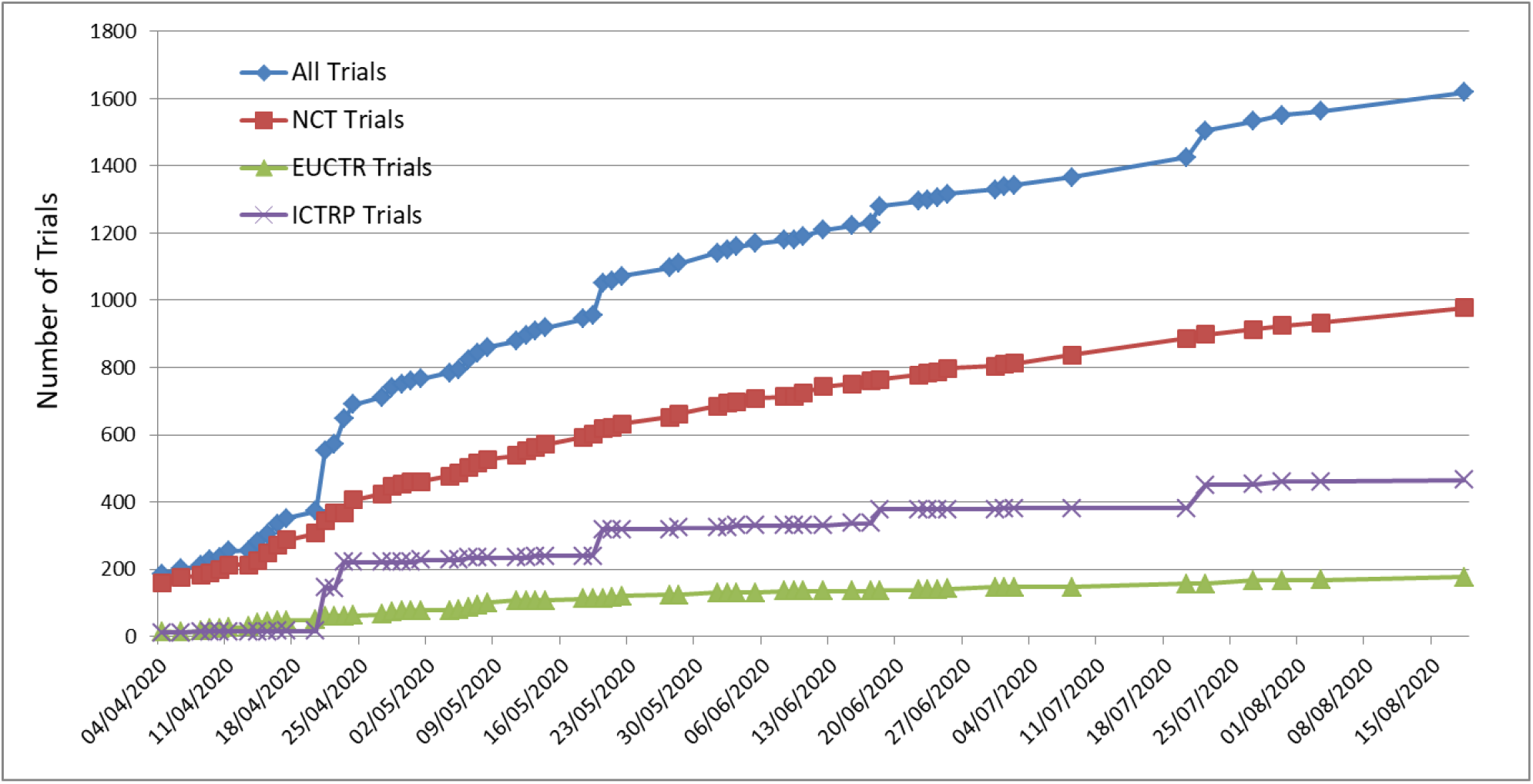
Number of trials from NCT, EUCTR and ICTRP included in covid19db.

The geographic distribution of trial activity has also expanded over the same period, as shown in Figure 3. Figure 3A shows the geographic distribution of trials as of May 19^th^. Figure 3B shows the increase in the number of countries with at least one in-scope COVID-19 trial registration, from 29 to 74 countries across the period.

**Figure 3.**
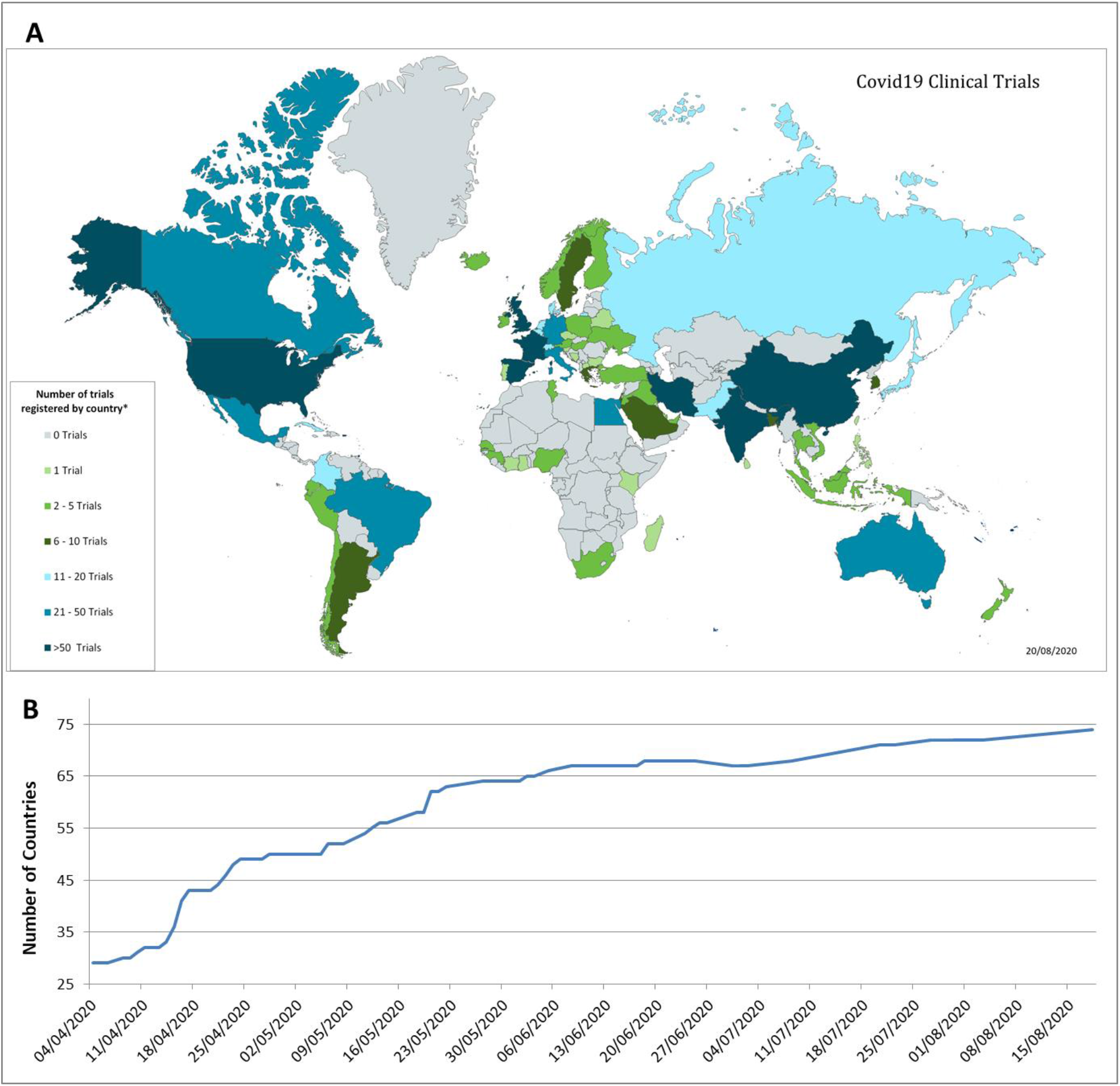
Geographic distribution of clinical trials.

The majority of trials, in both prevention and treatment settings, have included repurposing arms, as shown in Figure 4. While there has been an increase in the number of clinical trials for prevention of infection by SARS-CoV-2 over the period covered by this report, the proportion of trials for prevention has actually declined as a proportion of the total (from a maximum of 16% to 12.5% of the total).

**Figure 4.**
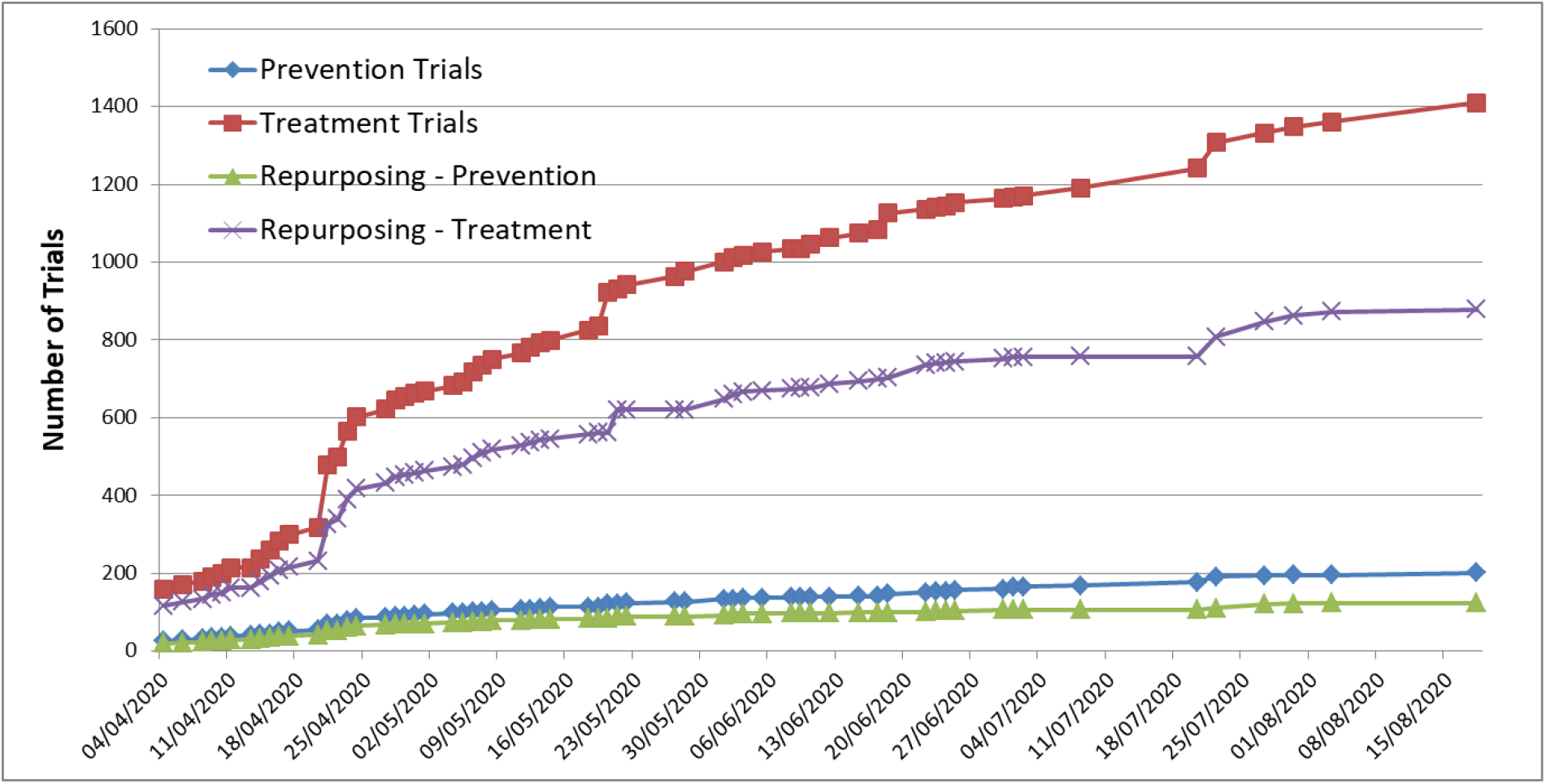
Number and proportion of clinical trials for treatment and prevention.

A notable feature of the response to the pandemic has been the wide range of interventions being tested in clinical trials, both in therapeutic and prevention settings. This is shown in Figure 5

**Figure 5.**
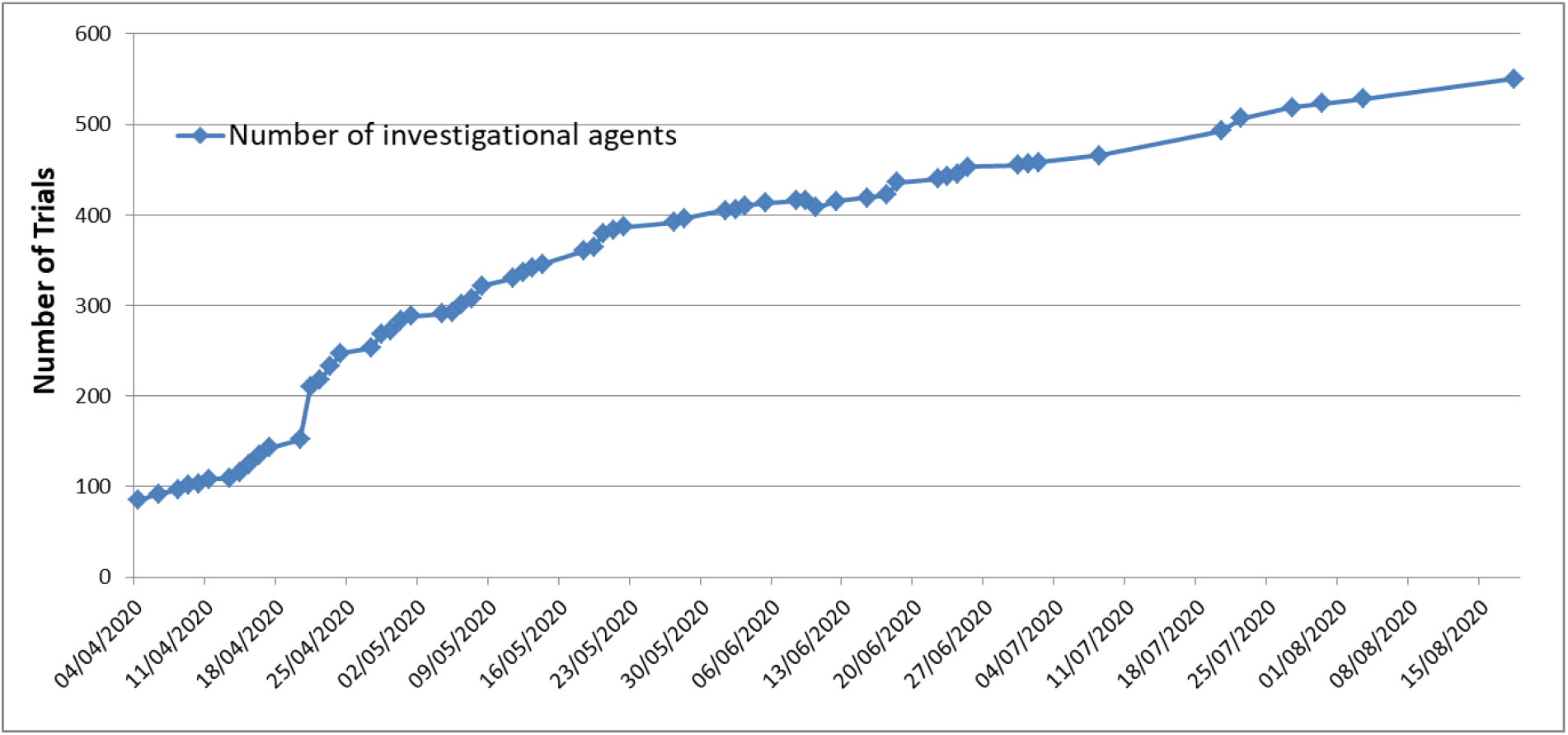
Increase in number of investigational agents included in trials.

The most prominent of the repurposed drug candidates was the anti-malarial/anti-rheumatic agent hydroxychloroquine – subject of much high-level political discussion and media attention. This agent is being trialled both for prevention and treatment, and in all patient populations. The increase in the number of trials of hydroxychloroquine is shown in Figure 6.

**Figure 6.**
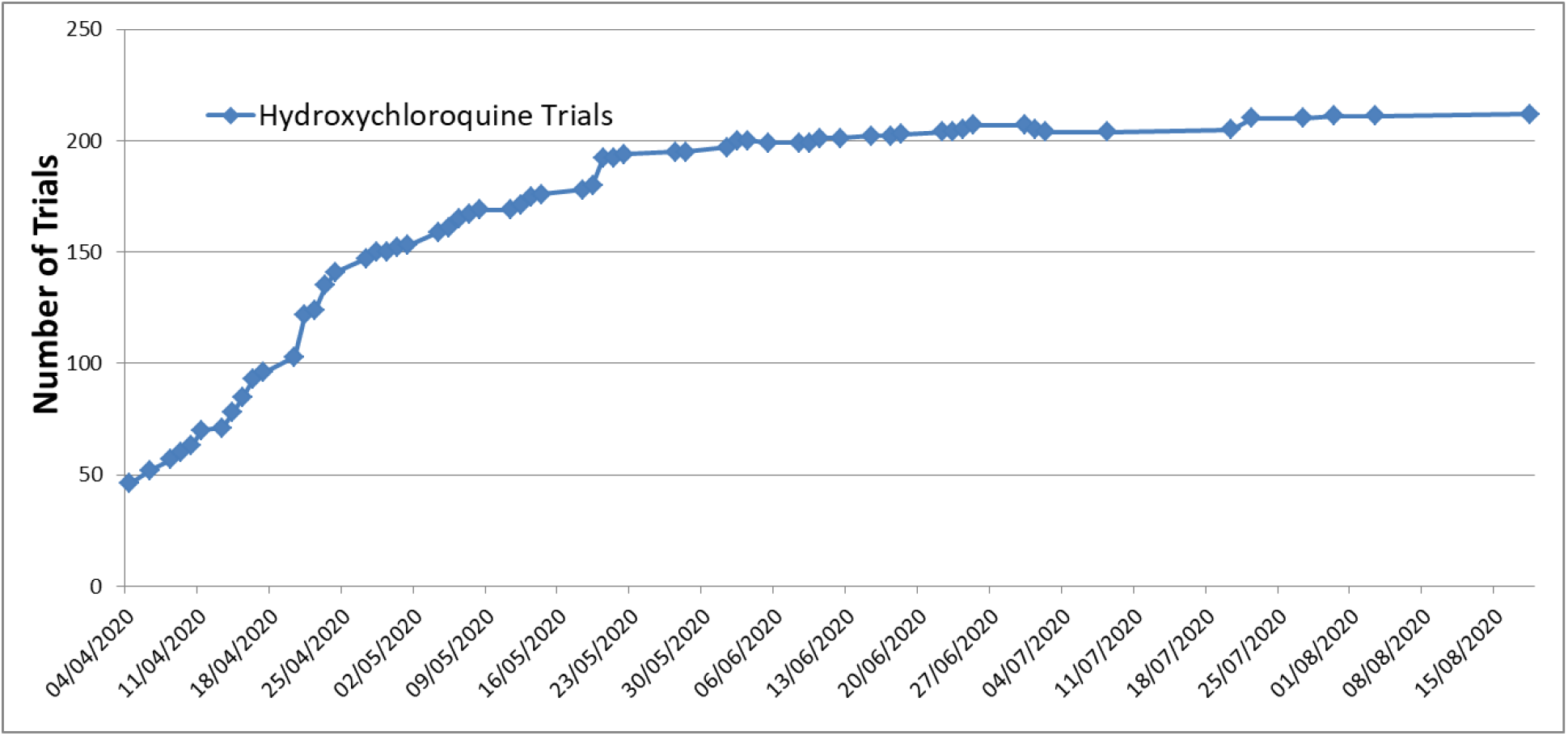
Number of trials including hydroxychloroquine as an investigational drug (both prevention and treatment trials)

The most popular drugs and types of intervention, as of August 18^th^, are shown in Table 1. Note that percentages of interventions sum to more than 100% as some trials may include more than one type of intervention.

**Table 1.** Most popular drugs and intervention types.

| Drug | n | % |
| --- | --- | --- |
| Hydroxychloroquine | 212 | 13% |
| Azithromycin | 64 | 4% |
| Tocilizumab | 58 | 4% |
| Favipiravir | 44 | 3% |
| Chloroquine | 43 | 3% |
| Lopinavir/ritonavir | 43 | 3% |
| Ivermectin | 39 | 2% |
| Methylprednisolone | 31 | 2% |
| Ascorbic acid | 30 | 2% |
| Intervention Type | n | % |
| Drug Repurposing | 1036 | 64% |
| Cell/Blood based | 279 | 17% |
| Shelved drug or NME | 267 | 17% |
| Other | 16 | 1% |

Finally, in terms of the planned recruitment, here too there was a sustained increase in numbers during the period covered in this report, as shown in Figure 7.

**Figure 7.**
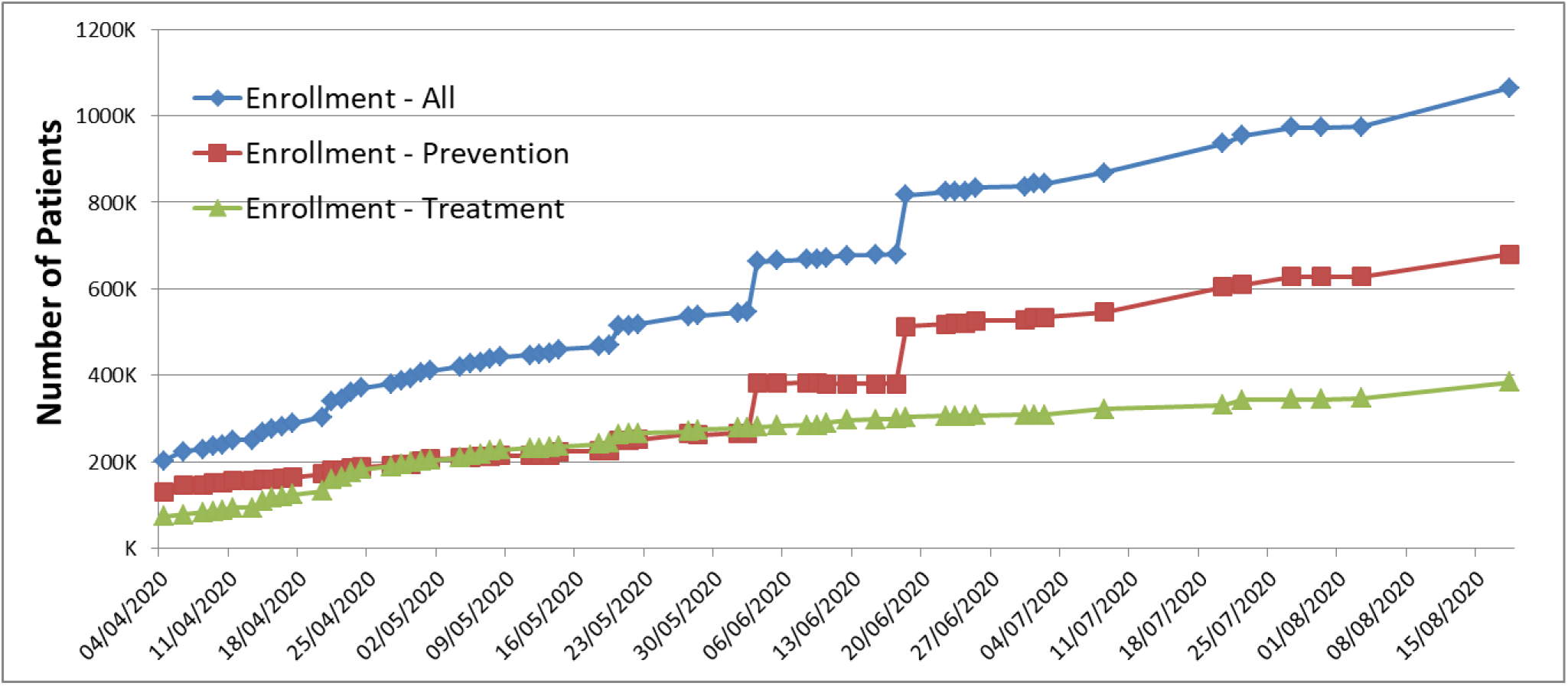
Planned enrolment for trials included in Covid19db.

## Discussion

We have developed a tool to monitor the registration of trials of medicinal products against COVID-19. Using this tool, we have confirmed that the majority of trials have included one or more repurposing arms – with a proportion of 64 – 75% across the period covered. Also confirmed is the very high number of trials investigating hydroxychloroquine. However, it is also notable that the number of different agents being investigated in trials has risen consistently – from fewer than in 100 in early April to 550 in mid-August 2020. It is likely that the proportion of repurposed agents has peaked and that as the pharmaceutical industry has brought new and shelved compounds to trial the proportion of repurposing trials has shown a slight decline.

The use of repurposing as a strategy has been striking during this period. In contrast to other areas of medicine, repurposing has been at the forefront of clinical trial activity for COVID-19 [3]. The rationale for this is clear – the availability of existing medicines and the speed with which these could be deployed in trials is a clear advantage. Many of the drugs being trialled show activity on relevant biological pathways and/or have been used previously in other viral diseases. Repurposing, therefore, is a rational response to the dire situation that has presented itself to the world. However, there are also many trials have been initiated based on scant evidence, such as *in vitro* or mechanistic data only [8]. The danger for those of us working in the field of drug repurposing is that such trials, and the huge attention paid to drugs such as hydroxychloroquine, threaten to discredit the idea of repurposing in the wider medical, research and lay communities.

We note that while the number of prevention trials was far lower than the number of treatment trials, planned enrolment for prevention trials is higher. It is also clear, as shown in Figure 3, that this is a global effort, with all parts of the globe hosting trials. Planned enrolment across all trials is well over one million patients and continues to grow. Whether such numbers are successfully recruited remains an open question. As the rate of infection declines it may well be that some trials will not meet recruitment targets and may have to be terminated, as is reported to have happened in China [9].

Our tool has some strengths and weaknesses. It is publicly available and the data are easily downloadable. Since we include the WHO ICTRP data, the EUCTR and clinicaltrials.gov, it provides global coverage of clinical trials activity of medicinal products against COVID-19. Our methodology also allows for rapid update as new trials are registered. Manual assessment of each trial has both advantages and shortcomings. Thanks to this manual assessment we have been able to eliminate duplicates and non-interventional trials, and to characterize interventions truly tested in trials (and not listed because of their use in the control group). However, the manual assessment may also lead to rare, though possible, coding errors. In the case of duplicate registrations only one record is used as the reference record and the duplicate entries discarded, therefore the counts for the individual registries, as shown in Figure 2, may not accurately reflect the true number of COVID-19 trials included in them.

Our work is however limited by the quality of the information included in the different registries. Common issues include unclear data regarding specific interventions, missing or incomplete data in key fields such as planned enrolment or number and location of trial sites, duplicate entries for some trials within a single registry. Data analysis is also complicated by the lack of API access to some registries – using web spidering technology is inherently brittle and inferior to full programmatic access to data. Finally, the lack of a universal identifier system for trials means depending on local identifiers, such as NCT number, rather than having an unambiguous method to identify unique trials.

## Conclusions

We may speculate on the influence of political interventions, the need to institute trials to gain access to drugs and other factors but it also remains true that such duplicated efforts ultimately do not serve science or patients [1,2]. However, there are also some very positive examples that have emerged from this crisis. The Solidarity trial, organised by the World Health Organisation, is a multi-national four-arm trial that has implemented a novel simplified web-based procedure to enable local institutions to participate [10,11]. Also notable is the Recovery trial from the UK [12]. The trial moved very rapidly from conception to approval to recruitment – in fact nine days from writing the protocol to recruitment of the first patient. Recruitment has been similarly impressive – as of 20th August 2020 12240 patients had been recruited in 176 centres. Preliminary results have highlighted the utility of dexamethasone in patients on invasive mechanical ventilation or oxygen without invasive mechanical ventilation [13]. The Discovery Trial, from France, is also a notable large, multi-arm trial with planned accrual across Europe.

At the other extreme there have been numerous small ‘me-too’ trials exploring the same range of drugs, with small sample sizes, lack of controls, or non-standard ‘standard of care’. The wide range of agents being explored also raises questions as to the levels of evidence required to support the rationale for some trials. Indeed, it has already been described as a torrent [14].

In addition to finding answers to the medical challenges imposed by SARS-CoV-2 it is hoped that the experience gained in the clinical trial arena, both positive and negative, may lead to improvements in trial design and regulatory processes that extend beyond this medical emergency [15].

## Data Availability

Data available online at http://www.redo-project.org/covid19db/

